# Examining the Impact of Chronic Pain on Information Processing Behavior: An Exploratory Eye-tracking Study

**DOI:** 10.1101/2022.02.15.22270955

**Authors:** Doaa Alrefaei, Gaayathri Sankar, Javad Norouzi Nia, Soussan Djamasbi, Diane Strong

## Abstract

Chronic pain is a multifaceted complex experience that is often captured with self-reported measures. While subjective self-reported measures capture pain from a patient’s point of view, they are limited in information richness. Collecting eye movements when completing self-reported subjective pain measures provides valuable insight about information processing and decision behavior. This information can improve the information richness of self-reported pain measures by providing a broader view of an individual’s pain experience. How people process information and make decisions when completing pain measures can also help to investigate the cognitive-evaluative aspects of chronic pain, which in turn can provide insight for developing eye-tracking biomarkers of chronic pain, and by doing so help develop smart clinician support technologies. Our preliminary results show that people with chronic pain expended significantly more cognitive effort than their pain-free counterparts when completing three self-reported pain measures that are widely used in clinical settings. These results are promising because they suggest that eye movements may serve as valuable information to accompany self-reported pain scores and thus enable effective assessment and management of chronic pain. The results also suggest that eye movements may serve as suitable biomarkers of chronic pain.

## 1 Introduction

Chronic pain, defined as pain that persists for at least three months (Merskey, 1986), is a major public health problem. In the United States, chronic pain is one of the most commonly experienced chronic conditions, afflicting about 50 million (1 out of 5) adults (CDC, 2020; Yong et al., 2021). Pain refers to “a distressing experience associated with actual or potential tissue damage with sensory, emotional, cognitive, and social components” (Crofford, 2015). Chronic pain occurs when that pain persists over months.

Chronic pain impacts quality of life by limiting work and life activities; it often negatively affects individuals’ physical and mental health as well as their family and social relationships. In addition to having a negative impact on quality of life, chronic pain also has negative economic costs including reduced productivity levels, increased compensatory payments due to disabilities caused by persistent pain, and increased health care costs and medical expenses (CDC, 2020; Phillips, 2006; Yong et al., 2021).

Effective treatment and management of chronic pain starts with comprehensive assessment of pain experience and impact. Currently, chronic pain is assessed by capturing self-reported level of pain intensity and level of interference that chronic pain has on one’s daily function (McCahon et al., 2005; Rose et al., 2018). Such self-reported measures are instrumental in capturing chronic pain experience from the patients’ point of view.

Self-reported measures, by their nature, provide only a narrow view of pain experience because they require patients to summarize a multifaceted complex experience into a single subjective score (Xu & Huang, 2020). Because they are subjective and lack rich information, they are limited in providing physicians with the information they need for effective treatment.

Information about how people go about providing such a score when completing a self-reported measure (i.e., how they process the provided information to decide which single score best represents their pain experience) may help to broaden the inherently narrow view of pain experience that is captured by such process. For example, capturing the type and range of information when a patient is choosing a response may help to reveal the extent (low-high boundaries) of pain experience by that patient. Similarly, identifying visual elements that received the most intense attention can reveal pieces of information that served as anchors for decision making. These insights gained from information processing and decision behavior when completing self-reported measures provide a more comprehensive and dynamic understanding of pain experience, which in turn can help develop guidelines for more effective pain assessment, management, and treatment.

In this study, we take a first step towards the larger project of building such smart clinician support technologies, by examining the differences in viewing behavior and patterns of people with and without chronic pain when they summarize their pain experience into single scores in three pain-related scales in the Patient-Reported Outcomes Measurement Information System (PROMIS) 29+ profile measure: pain intensity, pain interference, and physical function.

To examine viewing behavior and patterns of people summarizing their pain experience, we conduct an eye-tracking study. Eye-tracking research shows that machine learning engines using only eye movements can automatically and reliably detect whether a user is experiencing higher/lower cognitive load (Shojaeizadeh et al., 2019). These studies, which show the reliability and predictive quality of eye movements in detecting a user’s experience of cognitive load, suggest that other experiences, such as chronic pain, may also be reflected in eye movements.

Identifying eye movement behavior and/or patterns that are affected by chronic pain not only provides a more comprehensive picture of pain experience and the effect it has on cognition and decision behavior, but also can help identify ocular behavior that can serve as effective biomarkers for chronic pain. Such biomarkers can in turn be used in designing machine learning engines that can detect the presence and intensity of chronic pain in real-time to support health professionals in the development of personalized pain treatment and management solutions.

## 2 Background

Our approach to developing richer and more objective assessments of chronic pain is based on two results from the pain literature, that those experiencing pain differ in their allocation of attention to pain stimuli and they differ in their cognitive processes such as those involved in decision making. These results, which are briefly discussed below, basically suggest that pain affects how people process information and how they use that information to make decisions. Because eye-tracking provides unobtrusive insights into attention and cognition related to decision making, we conjecture that eye-tracking could be used to differentiate between those who are experiencing chronic pain and those who are pain free, and thus provide biomarkers of pain.

### 2.1 Pain and Attention

The pain literature suggests that allocation of attention to pain stimuli can reveal the presence of pain experience (Chan et al., 2020). For example, studies show that people in chronic pain often gravitate towards pain stimuli. This phenomenon is referred to as pain-related attentional bias and is a major focus of chronic pain literature. Pain-related attentional bias is typically studied via stimulus presentation methods that measure attention by reaction time to pain/non-pain pairs of stimuli, such as sensory words and images (Chan et al., 2020).

More recently, chronic pain studies have used eye tracking to directly measure how attention is allocated to such pairs of stimuli (Fashler & Katz, 2014; Franklin et al., 2019; Lee et al., 2018; Vervoort et al., 2013). These studies are grounded in the ‘‘eye-mind’’ assumption (Just & Carpenter, 1980), which has led to wide agreement that eye gaze serves as a reliable measure of attention. Eye gaze is measured with eye trackers, which record moment-to-moment ocular behavior, providing a continuous record for when, for how long, and how many times a person paid attention to a stimulus (Djamasbi, 2014).

### 2.2 Pain and Decision Making

Chronic pain affects cognitive processes (e.g., attention, perception, and evaluation) that are fundamental in judgment and decision making (Moriarty et al., 2011). Hence, chronic pain studies may benefit from extending their investigations from examining attentional biases to pain related stimulus to examining how stimuli are used to make pain related decisions. We take this approach in this study by examining the impact of chronic pain on information processing and decision behavior of individuals when they complete a pain measure.

Self-reported pain measures provide suitable stimuli for investigating how chronic pain impacts decision behavior. These subjective measures assess chronic pain by asking people to select the response that best summarizes their pain experience among a set of available responses. By doing so, these subjective measures provide an excellent opportunity for observing differences in attention to information that is needed for making judgments and decisions between people with or without chronic pain. Additionally, because self-reported pain measures are widely used in clinical settings, they provide an ecological valid paradigm of investigation that increases confidence in the generalizability of the results.

### 2.3 Pain Assessment with PROMIS

Through an initiative by the National Institutes of Health (NIH), PROMIS measures were developed to provide a standardized national resource for clinicians and scholars to assess and/or monitor an individual’s physical, mental, and social well-being (Cella et al., 2010; Rose et al., 2018). The PROMIS profile includes measures in a number of health domains such as physical function, anxiety, fatigue, depression, cognitive function, ability to participate in social roles, sleep disturbance, pain interference, and pain intensity. In this study, we used three measures from the PROMISE 29+ profile, namely pain intensity, pain interference, and physical function, because these are the most direct measures of chronic pain and the ones of initial concern by physicians. That is, the starting point in chronic pain assessment is evaluating the intensity of one’s pain, how much the pain interferes with one’s daily life, and how much the pain affects one’s physical functioning.

The pain interference and physical function measures used in our study, each included four items that were scored on a 5-point numeric rating scale. Larger scores for these two measures indicated higher levels of interference with daily activities and more difficulty with physical functioning. The pain intensity measure included one item that was scored on an 11-point numeric rating scale from 0-10, where 0 indicated no pain at all and 10 indicated worst pain imaginable.

In this study, we compare the information processing and decision behavior of people with and without chronic pain. To do so, we first categorized the data from individuals who participated in our study into two groups (chronic pain and pain free groups) based on participants’ own self-identification as suffering from chronic pain or being pain free.

In this study we use the pain intensity, pain interference, and physical function measures in two ways. First, we use the self-reported scores obtained from these measures to assess the differences in pain experience levels of participants in the chronic pain and pain free groups. This comparison allows us to see whether there were major differences in the degree to which these two groups experienced pain. Understanding the difference in pain experience between the groups could potentially help with interpreting the eye tracking results. The more pronounced the difference in pain experience between the two groups the more likely we will find differences in information processing and decision behavior in this initial step of our project.

In addition to using the scores of the self-reported pain intensity, pain interference, and physical function measures to assess differences in level of pain experience between the two groups, we use the items, and response scales of these measures as the stimuli for an eye-tracking experiment to assess whether eye gaze data, as indicators of visual information processing and decision behavior, differ for participants with and without chronic pain.

## 3 Methodology

Our study is an IRB-approved eye-tracking experiment using two groups of subjects, those who are pain-free and those with chronic pain. Participants in our study complete the pain intensity, pain interference, and physical function measures in the PROMISE 29+ profile while their eye movements are captured by an eye-tracking machine. The scores for the three aforementioned measures as well as the eye-tracking data captured while these measures were completed by study participants, were analyzed to produce the results reported in the next section.

### 3.1 Participants

Thirty-nine graduate and undergraduate students participated in our study which took place over a seven-week period. Each participant received a $20 gift card as a token of our appreciation. It is often the case that the eye tracker cannot be calibrated for a small percentage of participants in a study (Fehrenbacher & Djamasbi, 2017). In our experiment, we encountered one such case. Hence, the data for this participant was excluded from the analysis reported in this paper.

The remaining 38 sets of data were then grouped into chronic pain (n=12), pain free (n=22), and in-between (n=4) categories based on participants’ self-identification as suffering from chronic pain, being free of chronic pain, or being somewhere in-between these two conditions. Because in this study we were interested in comparing the ocular behavior of people with and without chronic pain, we removed the recordings for those four individuals who self-identified as “in-between” condition from the analysis. This process resulted in a dataset with gaze movement recordings for a total of 34 participants.

### 3.2 Task

To study information processing and decision behavior, we used three self-reported pain measures (i.e., pain intensity, pain interference, and physical function) as the visual stimuli in our study. The primary task in our study required participants to process the stimuli by providing responses to the items of these three self-reported measures. The task was presented to participants via a desktop computer while their eye movements were being captured unobtrusively by an eye-tracking machine attached to the monitor.

### 3.3 Data Collection Procedure

The eye movements of each participant were collected individually in the eye-tracking lab in a northeastern U.S. university. We used the Tobii Pro Spectrum 600HZ to collect participants’ eye movements. We used the IVT filter provided by Tobii Pro Lab software to identify fixations and saccades in the raw gaze stream. The IVT filter threshold was set on 30°/s and the minimum fixation duration was defined as 100ms (Liu et al., 2021).

After calibrating the eye tracker for participants, they were asked to complete the task (complete the three pain self-reported measures). After participants competed the task, we conducted an exit interview, during which we provided participants with the definition of chronic pain and asked them to self-identity their pain status.

### 3.4 Measures

We used four subjective self-report and four objective eye-tracking measures in our study. One of the subjective measures was used to categorize participants into two pain-status (chronic pain and pain free) groups. The other three subjective measures were used to determine the degree to which the two groups differed in pain experience. These measures are explained below.

#### Subjective pain status (1 measure)

We used this measure to categorize participants into chronic pain and pain free groups. We provided participants with the definition of chronic pain, i.e., a pain experience that is rated as 4 or higher on a 0-10 low to high scale and lasts more than 3 months. Then, we asked participants to tell us how they would describe their pain status: would they self-identify as 1) someone who is suffering from chronic pain, 2) someone who is pain free, 3) or someone who has a pain experience that is somewhat in-between the chronic pain and pain free conditions. Only the data for chronic pain and pain free groups were used in the analysis reported in this study.

#### Subjective pain measure (3 measures)

The experimental task in our study required participants to complete three subjective pain measures. In addition to using these pain measures as visual stimuli for the task, we used their ratings to investigate the degree to which participants in the chronic pain and pain free groups differed in pain experience. For example, we used ratings for the pain intensity measure to examine the differences in the severity of experienced pain between the chronic pain and pain free groups. Similarly, we used ratings for the pain interference and the physical function measures to assess differences between the two groups in the degree to which pain interfered with their daily life and the degree to which pain impacted their physical function.

#### Objective measures of eye movements (4 measures)

We computed three quantitative metrics for eye movement behavior that capture attention: fixation count, fixation duration, and visit count. Fixation count refers to the number of moments when the eye remains still on a stimulus. Fixation duration refers to total processing time for viewing a stimulus. Visit count refers to the number of times a given stimulus is visited by a viewer.

We used the relative fixation duration heatmap, a qualitative measure of attention, to explore differences in viewing patterns, e.g., the dispersion of fixations on various parts of a stimulus. Because eye-tracking heatmaps overlay the aggregated gaze data on stimulus, they provide an excellent tool for detecting patterns of visual attention. In addition to patterns, heatmaps reveal intensity of attention by using colors (e.g., using red, yellow, and green to visualize high to low fixation intensity). The relative fixation duration heatmap that we use in our analysis, depicts the intensity of each participant’s attention to various areas of a stimulus relative to the participant’s total attention to the entire stimulus.

### 3.5 Analysis

The subjective self-identified pain status (chronic pain”, “pain free”, or “in-between”) will be used to group datasets into chronic pain and pain free categories. The data for participants who self-identified as having an “in-between” pain status will not be included in the analysis in this study.

We will use the scores that were obtained from the self-reported pain measures during the experimental task (i.e., subjective ratings for pain intensity, pain interference, and physical function) to test whether differences in pain experience between participants in the chronic pain and pain free groups is significant or not. Because pain affects cognition, lack or presence of significant differences in pain experience can provide better explanations for the eye-tracking results. For example, the more pronounced the differences in pain experience between the two groups the more likely we will observe differences in information processing and decision behavior of participants with and without chronic pain.

For eye movement analysis, as customary in eye-tracking research (Djamasbi, 2014), we will define areas of investigations or interest (AOIs) that are relevant to the visual stimuli used in our study. The task in our study requires participants to complete three self-reported pain measures, each of which requires participants to read a set of pain related questions and respond to those questions by selecting an option on a numeric scale. Hence, visual stimuli used in our study contains three essential AOIs. The first AOI is the part of the stimulus, or self-reported measure, that contains the questions (Question AOI) to be answered. This question region is separate from the answer region of the stimulus that contains possible answers to those questions. The second and third AOIs separate the answer region on the stimulus into two separate AOIs: one that contains the labels for the numeric scale (Label AOI) and one that contains the response options (Response AOI). We will use the quantitative and qualitative eye tracking metrics explained in the previous section to compare differences in viewing behavior and patterns in the AOIs between the two groups.

## 4 Results

### 4.1 Findings from Self-reported Measures

To prepare the datasets for analysis, we grouped them into chronic pain and pain free groups based on participants’ self-reported pain status, i.e., participant’s self-identification as suffering from chronic pain or being free of chronic pain. Next, we compared participants’ scores for the three self-reported pain measures (pain intensity, pain interference, physical function) between the two groups. Participants in the chronic pain group provided significantly (p < 0.01) higher scores for pain intensity (5.42 vs. 0.68), pain interference in daily function (2.67 vs. 1.12), and difficulty in physical function (1.79 vs. 1.10). Based on these results, which are displayed in Figure 1, we conclude that participants in the chronic pain group had a significantly heightened pain experience as compared to participants in the pain free group.

**Fig. 1.**
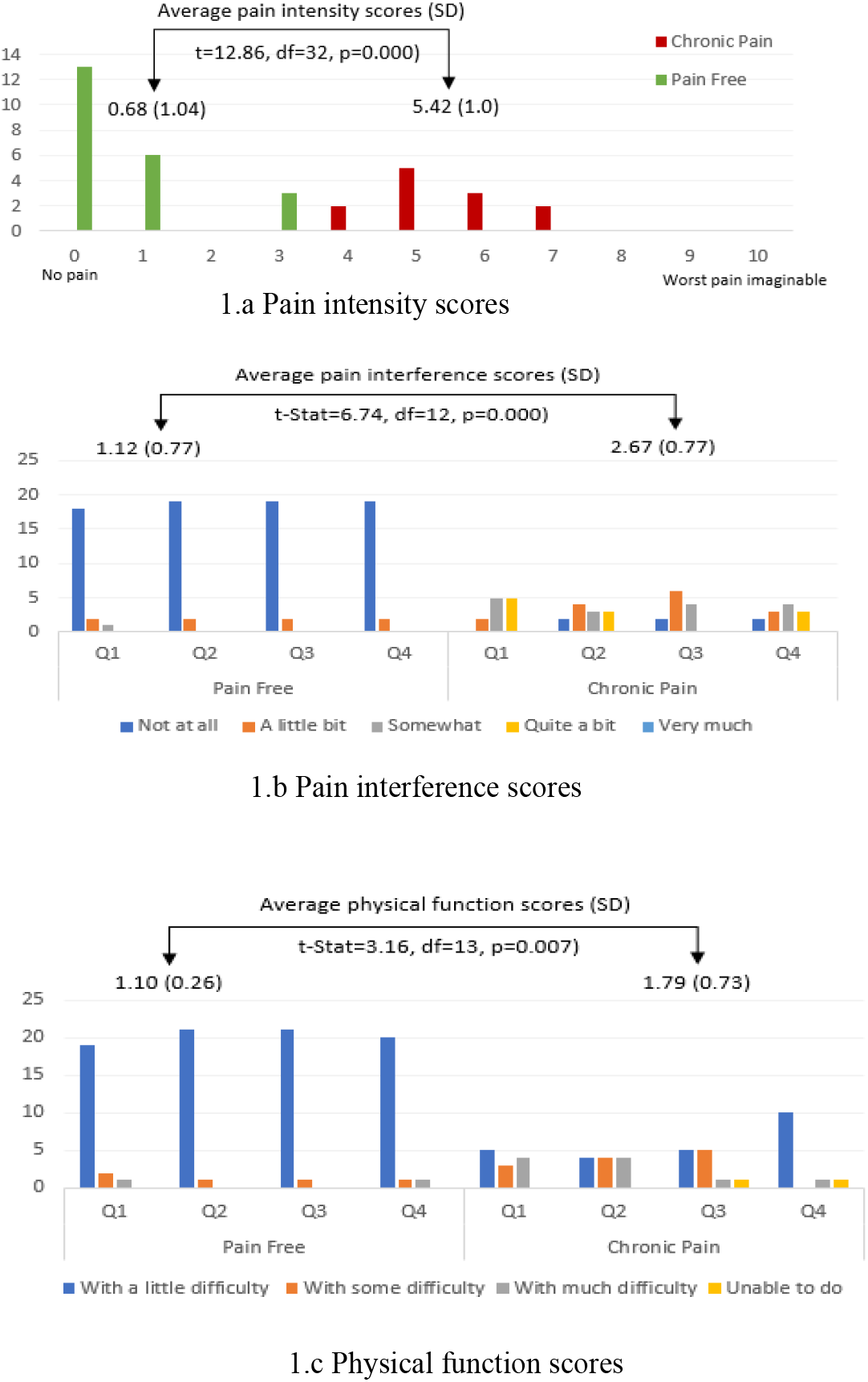
Ratings for pain experience self-reported measures.

Next, we analyzed the eye movement data captured during the completion of the three pain measures (i.e., pain intensity, pain interference, and physical function measures) to examine differences in information processing and decision behavior between the two groups.

### 4.2 Findings from Eye Movement Metrics

The eye movement analysis in this section is organized in three separate parts, each reporting the results for one of the three stimuli, i.e., pain intensity, pain interference, and physical measures. The results reported in each part are summarized by a table that is organized first by eye movement metrics and then by the defined AOIs. Each part also provides a heatmap for the analysis of fixation patterns within the defined AOIs. Fixation patterns on these heatmaps are visualized with colors red, yellow, and green representing high, medium, and low levels of fixation intensity. The combination of quantitative and qualitative eye tracking data has been shown to extend the explanatory power of eye tracking analysis (Djamasbi et al., 2011).

#### Pain Intensity

The results of t-tests did not show significant differences between the two groups in fixation and visit metrics for the Question AOI on this stimulus. There were also no significant differences between the groups in fixation durations in any of the AOIs. The results, however, showed that people in the chronic pain group had significantly (p=0.04) fewer fixations (3.58 vs. 6.36) in the Label AOI and visited this AOI significantly (p=0.01) less frequently (1.92 vs. 3.14) than people in the pain free group. People in the chronic pain group had almost significantly (p=0.07) more fixations in the Response AOI (Table 1).

**Table 1.** t-test results for the pain intensity measure

| Fixation Count | Pain Free | Chronic Pain |  |
| --- | --- | --- | --- |
| Question AOI | 6.09 (3.84) | 6.00 (2.49) | $t=0.08$, $df=32$, $p=0.94$ |
| Response AOI | 7.5 (3.83) | 10.58(5.57) | $t=1.90$, $df=32$, $p=0.07^*$ |
| Label AOI | 6.36 (4.09) | 3.58 (2.81) | **$t=2.09$, $df=32$, $p=0.04$** |
| Fixation Duration (ms) |  |  |  |
| Question AOI | 1256.41 (1010.26) | 1130.92 (475.60) | $t=0.49$, $df=32$, $p=0.62$ |
| Response AOI | 2356.96 (1596.68) | 3135.42 (1794.32) | $t=1.30$, $df=32$, $p=0.20$ |
| Label AOI | 1177.14 (823.72) | 756 (833.15) | $t=1.41$, $df=32$, $p=0.17$ |
| Visit Count |  |  |  |
| Question AOI | 2.36 (1.62) | 2.00 (0.60) | $t=0.75$, $df=32$, $p=0.46$ |
| Response AOI | 3.14 (1.13) | 3.5 (1.00) | $t=0.93$, $df=32$, $p=0.36$ |
| Label AOI | 3.14 (1.32) | 1.92 (1.24) | **$t=2.63$, $df=32$, $p=0.01$** |

\*Almost significant

Viewing patterns shown in the Question AOI were dispersed covering the entirety of the textual information (Figure 2). This behavior represents careful processing of text-based communication (Djamasbi et al., 2011). Viewing patterns in the Response and Label AOIs, which supported the results displayed in Table 1, show that the two groups differed in how their fixations were distributed when they were deciding on a response to represent their experience. People in the chronic pain group looked at the last 8 last numeric options in the Response AOI to make decisions. Even though no one in the chronic pain group selected responses 3, 8, and 10 (see Figure 1.a), these response options received relatively intense fixations (Figure 2a). People in the pain free group looked at the first 6 and the last 2 response options (Figure 2.b) while the majority of participants in this group chose the first two responses (Figure 1.a). Another notable difference in fixation patterns between the two groups is the way they viewed the Label AOI. Fixation patterns in Figure 2 show that people in the chronic pain group considered only the “worst pain imaginable” label when trying to assess their pain experience while people in the pain free group used both labels to make the same decision.

**Fig. 2.**
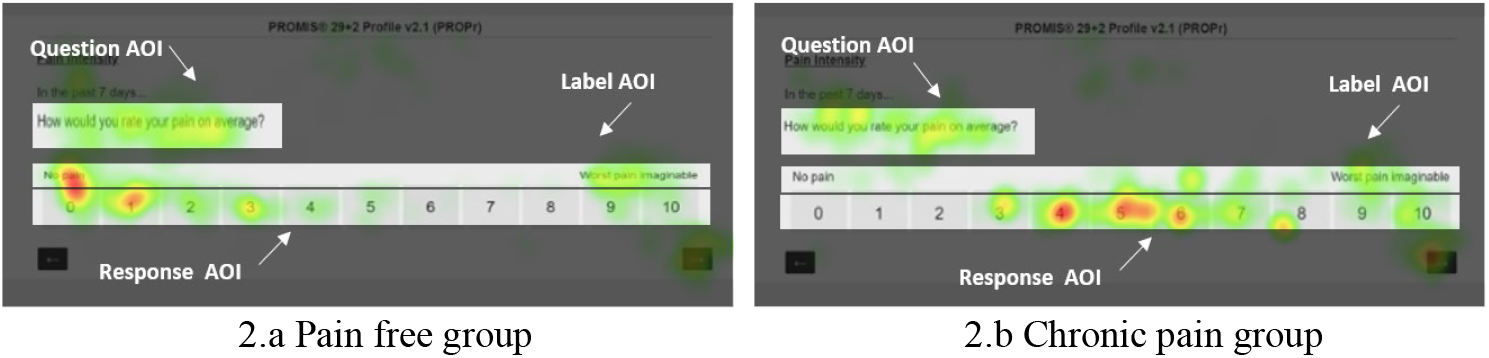
Relative fixation duration heatmaps for pain intensity.

These results together show that people in the pain group summarized their pain experience into a single value by gauging how their experience faired against “worst pain imaginable”. They processed the Label AOI with significantly less effort than the pain free group, but processed the Response AOI with almost significantly (p=0.07) more effort than the pain free group.

#### Pain Interference

The results of t-tests were not significant for differences in fixation/visit intensity between the two groups in the Question and Label AOIs for this self-reported measure. The intensity of fixations between the two groups, however, was significantly different when participants were processing the Response AOI of the pain interference measure. People in the chronic pain group had significantly (p=0.000) more (15.92 vs. 8.77) and longer fixations (3919.92 ms vs. 2144.36 ms) in this AOI and visited it significantly (p=0.002) more frequently (8.17 vs. 5.55) than people in the pain free group (Table 2).

**Table 2.** t-test results for the pain interference measure

| Fixation Count | Pain Free | Chronic Pain |  |
| --- | --- | --- | --- |
| Question AOI | 38.77 (1689) | 38.5 (13.14) | $t=-0.05$, $df=32$, $p=0.96$ |
| Response AOI | 8.77 (3.94) | 15.92 (3.20) | **$t=5.38$, $df=32$, $p=0.000$** |
| Label AOI | 5.2 (3.02) | 7 (3.41) | $t=1.51$, $df=29$, $p=0.14$ |
| **Fixation Duration (ms)** |  |  |  |
| Question AOI | 7659.73 (4085.30) | 7422.92 (3094.07) | $t=0.17$, $df=32$, $p=0.86$ |
| Response AOI | 2144.36 (1123.06) | 3919.92(1252.42) | **$t=4.23$, $df=32$, $p=0.000$** |
| Label AOI | 1245.3 (884.97) | 1593.91 (806.04) | $t=1.08$, $df=29$, $p=0.29$ |
| **Visit Count** |  |  |  |
| Question AOI | 5.91 (1.69) | 6.00 (1.41) | $t=0.16$, $df=32$, $p=0.88$ |
| Response AOI | 5.55 (2.48) | 8.17 (1.53) | **$t=3.32$, $df=32$, $p=0.002$** |
| Label AOI | 2.77 (1.77) | 3.92 (2.23) | $t=1.64$, $df=32$, $p=0.11$ |

**Table 3.** t-test results for the physical function survey

| Fixation Count | Pain Free | Chronic Pain |  |
| --- | --- | --- | --- |
| Question AOI | 39.27 (17.06) | 39.67 (12.94) | $t=0.07$, $df=32$, $p=0.95$ |
| Response AOI | 9.23(3.60) | 12.42 (5.50) | **$t=2.04$, $df=32$, $p=0.049$** |
| Label AOI | 17.5 (9.88) | 23.83 (10.72) | $t=1.73$, $df=3$, $p=0.09$ |
| **Fixation Duration (ms)** |  |  |  |
| Question AOI | 7775.64(4111.79) | 7297.58(2505.20) | $t=0.37$, $df=32$, $p=0.72$ |
| Response AOI | 2332.86 (1011.83) | 2912.67(1348.79) | $t=1.42$, $df=32$, $p=0.17$ |
| Label AOI | 4204.96 (2990.81) | 5219.25(2193.48) | $t=1.03$, $df=32$, $p=0.31$ |
| **Visit Count** |  |  |  |
| Question AOI | 6.55 (2.60) | 7.33 (2.31) | $t=0.88$, $df=32$, $p=0.39$ |
| Response AOI | 6.32 (2.50) | 8.83 (3.43) | **$t=2.46$, $df=32$, $p=0.02$** |
| Label AOI | 5.64 (3.03) | 7.33 (3.17) | $t=1.54$, $df=32$, $p=0.14$ |

Figure 3 displays the relative fixation duration heatmaps for the pain interference measure. These heatmaps show that both groups exhibited similar thorough processing patterns for the Question AOI evidenced by the dispersed fixation patterns covering the entirety of the textual information in this area. The heatmaps, however, show that the two groups had notable differences in viewing patterns in the Response and Label AOIs. People in the chronic pain group had more dispersed fixations in the Response AOI than people in the pain free group (evidenced by larger colorful areas). They also showed a more dispersed viewing pattern in the Label AOI. People in the chronic pain group looked at more labels, in particular the first four labels, to choose a response while people in the pain free group viewed the first three labels for decision making. The most intense fixations of people in the chronic pain group were on labels “a little bit”, “somewhat”, and “quite a bit”. The most intense fixations of people in the pain free group were on labels “not at all” and “a little bit”. These results reveal labels that were mostly considered for decision making by each group.

**Fig. 3.**
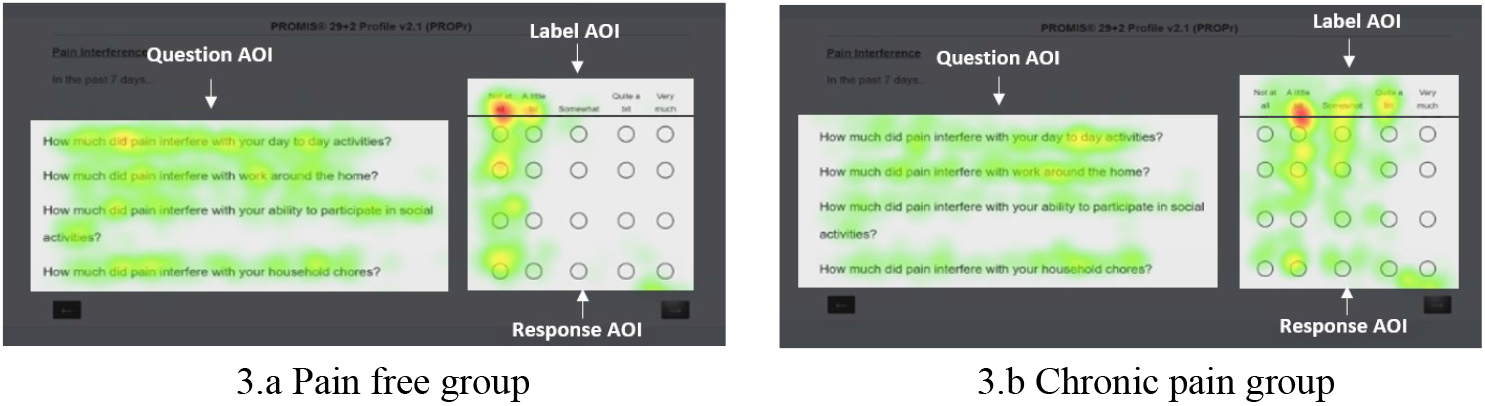
Relative fixation duration heatmaps for pain inerference.

The results displayed in Table 2 and the more dispersed pattern of fixations in the heatmap in Figure 3.a vs. the heatmap in Figure 3.b, together show that people in the chronic pain group expended significantly more cognitive effort to decide which response to choose.

#### Physical Function Stimulus

Once again, we found no significant differences between the two groups in how intensely they processed or how frequently they visited the Question AOI when completing the physical function measure. While we found no significant differences between the two groups in fixation and visit metrics in the Label AOI, our analysis showed that the chronic pain group processed the Response AOI with significantly (p=0.049) more fixations (12.42 vs. 9.23) and visited this AOI significantly (p=0.02) more frequently (8.83 vs. 6.32) than the pain free group.

Figure 4 displays the relative fixation duration heatmaps for the chronic pain and pain free groups when they were completing the physical function measure. As in previous heatmaps displayed in Figures 2 and 3, the heatmaps in Figure 4 show that both groups processed the Questions AOI with similarly dispersed fixation patterns that is representative of thorough information processing behavior (Djamasbi et al., 2011). Both groups viewed all of the labels in the Label AOI, but people in chronic pain had more intense fixations on the first four labels (“without any difficulty”, “with a little difficulty”, “with some difficulty”, “with much difficulty”). The most intense fixation of people in the pain free group was on the first label (“without any difficulty”). Fixation patterns of people in the chronic pain group were more dispersed covering the first four columns in the Response AOI. Fixation patterns of people in the pain free group were more focused covering mainly the first column of the Response AOI (Figure 4).

**Fig. 4.**
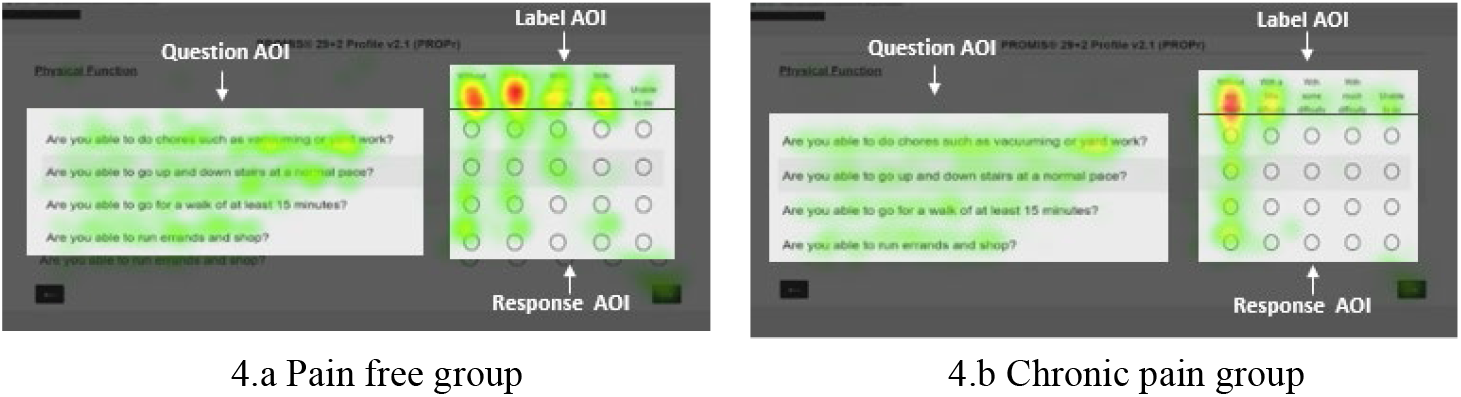
Relative fixation duration heatmaps for physical function.

The significantly higher number of fixations and visits in the Response AOI, along with the more dispersed fixation patterns in the Response AOI and having more intense fixation on a larger number of labels (Figure 4), show that people in the chronic pain group went through a more cognitively effortful decision process than those in the pain free group.

## 5 Discussion

We used eye tracking to compare information processing and decision behavior of people with and without chronic pain when they were summarizing their pain experience into a single score via responding to three pain related measures that are included in the PROMIS 29+ profile. We investigated information processing and decision behavior by examining attention on three complementary AOIs that were necessary for decision making: 1) Question AOIs delineating questions in pain measures, 2) Label AOIs labeling response options (e.g., “not at all”, “quite a bit”, etc.), and 3) Response AOI delineating the response options that were available for selection. Differences in attention to these AOIs between the two groups were determined by comparing how many times an AOI was viewed (visit count), how many fixations (fixation count) were used to process it, and how long those fixations lasted (fixation duration) in each AOI. To increase the explanatory power of this preliminary analysis, we used relative fixation duration heatmaps to examine fixation patterns and intensity on various parts of the AOI.

The qualitative and quantitative analysis of eye movements showed that questions in all three self-reported pain measures were processed thoroughly by both groups. No significant differences were detected between the two groups when they were reading the questions in the aforementioned pain measures (processing Question AOIs). The fixation pattern in both groups exhibited a thorough reading behavior by showing that fixations covered the entire text in the Question AOI.

When summarizing pain experience into scores along the given scales, however, our results showed that people with and without chronic pain had major differences in information processing and decision behavior. For example, the chronic pain group exhibited almost significantly more fixations than the pain free group in the Response AOI of the pain intensity measure. When making decisions about pain interference, the chronic pain group, significantly more than the pain free group, expended cognitive effort to process the Response AOI as evidenced by their significantly more visits and significantly more and longer fixations on this AOI. Similarly, the chronic pain group exhibited a higher level of cognitive effort when processing the Response AOI of the physical function measure by exhibiting significantly more frequent visits and significantly more fixations on this AOI than the pain free group.

The higher level of cognitive effort in the chronic pain group when responding to questions was also evidenced by their eye movement patterns. For example, the relative pain intensity heatmaps developed for the chronic pain group showed more red-colored clusters in the Response/Label AOI than those developed for the pain free group. Similarly, the number of red and bright yellow clusters in heatmaps for pain interference and physical function measures showed that the pain group had more relative intense fixations in the Response and Label AOIs than the pain free group. In these heatmaps, fixations of the chronic pain group covered a larger area of the Response/Label AOI than fixations of the pain free group, which is yet another indication of expending more cognitive effort (Djamasbi et al., 2011).

Heatmaps also revealed which labels were more heavily used in selecting responses. For example, the heatmaps in Figure 2 show that the chronic pain summarized their experience of pain intensity by focusing mainly on the “worst pain imaginable” label, while the pain free group considered both “no pain” and “worst pain imaginable” labels. The heatmaps for the pain interference measure (Figure 3) show that the pain group viewed the labels in the middle range of spectrum (labels in 2^nd^, 3^rd^, and 4^th^ place) more intensely than labels in the outer edges of the spectrum. The pain free group more intensely focused on the low end of the spectrum (the first two labels). The viewing pattern for the physical function measure (Figure 4) shows that both groups viewed all five labels. These heatmaps, however, show that the pain group fixated more intensely on the first four labels while the pain free group focused more on the first two labels. The pattern of focus on labels not only suggests differences in cognitive effort between the two groups (the more labels used to choose responses, the more effortful the decision making process) but also reveals notable differences in low-high range of label boundaries that they were used for decision by the two groups (e.g., “a little bit” to “quite a bit” vs. “not at all” and “a little bit” in Figure 3).

These differences can provide a more comprehensive picture of pain experience to help assess how intensely a person suffers from chronic pain. It can also potentially be used in developing objective markers of chronic pain. For example, Figure 2 shows that the pain group visited the label area significantly less frequently, with significantly fewer fixations, and focused only on the “worst pain experience” label when deciding which response best represented their pain intensity. The narrowly focused (using only one label) and decisive decision process (fewer fixations on the label) represents the pain experience reality of people who live with chronic pain. Suffering from chronic pain naturally excludes “no pain” from one’s experience palette. This interpretation also explains the fixation patterns of the pain free group for the same measure. While the pain free group viewed both labels of the pain intensity measure, the red color on the “no pain” label (Figure 2.b) suggests that this label, which is representative of their dominant pain experience, had a larger weight in their decision making.

The observed differences in information processing and decision behavior of people with and without chronic pain in our study suggest that eye movements may serve as reliable objective (physiological) biomarkers for chronic pain. Our results, showing the sensitivity of eye movement data in detecting differences in cognitive effort unobtrusively without additional burden to users, are promising for developing eye tracking machine learning engines that can detect in real-time whether and how intensely a person suffers from chronic pain.

Our results also suggest that the insight gained from eye movements may help provide more personalized interventions. Pain is a complex phenomenon that benefits from multi-dimensional assessments (van Boekel et al., 2017). Augmenting self-reported pain measures with the insight revealed by the objective eye movement data (e.g., the range of labels and anchors used in decision making) may help develop guidelines that more successfully can meet the unique needs of individual patients for pain treatment and/or management (NSW, 2021).

Advances in eye-tracking technology increasingly and positively affect the affordability of high-quality eye trackers (Djamasbi, 2014). This trend makes it increasingly realistic and practical to collect eye-movement data when patients complete pain measures at clinics or office visits. The additional insight provided by eye movements when patients complete pain measures can be coded along the self-reported scores in a user-friendly decision support feedback for practitioners in real-time. This feedback in turn can help practitioners gain a more comprehensive view of their patients’ pain experience and thus be able to have more effective visits with their patients and develop more successful treatment options for them.

### 5.1 Theoretical and practical implications

From a theoretical point of view, our study contributes to human computer interaction literature in two ways: 1) by examining a user attribute (chronic pain) that may affect how decision support systems are used (Djamasbi, 2007) and 2) by examining eye movement behavior that can contribute to developing smart user-centered systems for supporting chronic pain healthcare professionals. Our study also contributes to chronic pain literature in two ways: 1) by investigating the potential of eye movements as a biomarker for chronic pain and 2) by using the more context rich and ecologically valid decision-making paradigm of pain measures to study information processing and decision behavior.

From a practical point of view, our results show that collecting eye movements during completion of pain measures at clinics can augment the self-reported pain scores with user-friendly visual report (e.g., fixation duration heatmap) and/or coded visual feedback developed from eye-movement data (Jain et al., 2020; Jain & Djamasbi, 2019). Such augmented self-reported measures can provide clinicians with a more comprehensive picture of pain experience, which in turn can lead to more successful office visits and more effective personalized chronic pain treatment solutions. Advances in video-based eye tracking make it possible to capture eye movements unobtrusively and without any additional burden on users. Eye trackers can easily be attached to computer monitors, laptops, kiosks, and other digital screens. Hence, they can easily be added to computerized systems that are used for administering self-reported pain measures at clinics typically immediately before an office visit.

As more and more high-quality eye-tracking technologies are embedded in consumer grade laptops and mobile devices, such information can also be collected during remote office visits which sometimes are the only practical and/or cost-effective option for clinicians to treat their patients (e.g., during pandemics, natural disasters, and/or patients’ mobility issues).

### 5.2 Limitations and future studies

As with any experiment, our study has limitations. For example, we had a relatively small sample size. While small sample sizes are common in exploratory eye-tracking research, future studies with larger sample sizes may help to detect more significant differences in information processing behavior of people with and without chronic pain. The significant results obtained in our study, however, support the sensitivity and potential power of eye movements in detecting user experience (Shojaeizadeh et al. 2019), which is promising for developing eye tracking biomarkers of chronic pain.

The analysis of viewing behavior in our study was limited to three AOIs reflecting three major components of the pain intensity, pain interference, and physical function measures. Future studies can refine this analysis by creating more AIOs, e.g., a separate AOI for each question and its respective response area. Our preliminary analysis mainly focused on fixation and visit metrics, using saccadic and/or pupillometry metrics may provide additional insight.

## 6 Conclusion

The analysis of eye-tracking data showed differences in information processing and decision behavior for people with and without chronic pain when they were responding to three self-reported pain-related measures. The results showed that people with chronic pain, compared to those who did not suffer from chronic pain, exhibited eye movement behaviors and patterns that were representative of expending more cognitive effort. These preliminary results are promising because they suggest that eye movements may serve as suitable biomarkers of chronic pain, which can help in developing systems that can detect pain experience automatically and unobtrusively. The results also suggest that collecting eye movements when completing a clinical pain measure provides additional useful information that can help practitioners have a more comprehensive view of their patients’ pain experience.

## Data Availability

Data is not availble.

